# Investigation of genomic and transcriptomic risk factors in clopidogrel response in African Americans

**DOI:** 10.1101/2023.12.05.23299140

**Authors:** Guang Yang, Cristina Alarcon, Catherine Chanfreau, Norman H. Lee, Paula Friedman, Edith Nutescu, Matthew Tuck, Travis O’Brien, Li Gong, Teri E. Klein, Kyong-Mi Chang, Philip S. Tsao, David O. Meltzer, Million Veteran Program, Sony Tuteja, Minoli A Perera

## Abstract

Clopidogrel, an anti-platelet drug, used to prevent thrombosis after percutaneous coronary intervention. Clopidogrel resistance results in recurring ischemic episodes, with African Americans suffering disproportionately. The aim of this study was to identify biomarkers of clopidogrel resistance in African American patients.

We conducted a genome-wide association study, including local ancestry adjustment, in 141 African Americans on clopidogrel to identify associations with high on-treatment platelet reactivity (HTPR). We validated genome-wide and suggestive hits in an independent cohort of African American clopidogrel patients (N = 823) from the Million Veteran’s Program (MVP) along with *in vitro* functional follow up. We performed differential gene expression (DGE) analysis in whole blood with functional follow-up in MEG-01 cells.

We identified rs7807369, within thrombospondin 7A (*THSD7A),* as significantly associated with increasing risk of HTPR (p = 4.56 x 10^−9^). Higher *THSD7A* expression was associated with HTPR in an independent gene expression cohort of clopidogrel treated patients (p = 0.004) and supported by increased gene expression on *THSD7A* in primary human endothelial cells carrying the risk haplotype. Two SNPs (rs1149515 and rs191786) were validated in the MVP cohort. DGE analysis identified an association with decreased *LAIR1* expression to HTPR. *LAIR1* knockdown in a MEG-01 cells resulted in increased expression of SYK and AKT1, suggesting an inhibitory role of *LAIR1* in the Glycoprotein VI pathway. Notably, the *CYP2C19* variants showed no association with clopidogrel response in the discovery or MVP cohorts.

In summary, these finding suggest that other variants outside of *CYP2C19* star alleles play an important role in clopidogrel response in African Americans.

**Key Points:** SNPs within *THSD7A* were associated to clopidogrel resistance in African Americans.

*THSD7A* gene expression is higher in patients with clopidogrel resistance and in endothelial cells carrying the risk variants.

No association to high on therapy platelet reactivity or major adverse cardiac events was seen with *CYP2C19* star alleles in either the African American cohort investigated.

Decreased *LAIR1* gene expression in blood was associated to risk of HTPR and showed downstream effect on SKY and AKT.

## Introduction

Clopidogrel is an anti-platelet agent commonly prescribed to prevent ischemic events in patients with coronary artery disease (CAD), acute coronary syndrome (ACS), and in patients undergoing percutaneous coronary intervention (PCI)^1^. However, the response to clopidogrel can vary widely between individuals, with both pharmacodynamic response (as measured by P2Y12 reaction units [PRU]) or clinical response showing significant interindividual differences^2^. High on-treatment platelet reactivity (HTPR),^3^ has been linked to a greater incidence of major adverse cardiovascular events (MACE) in clinical trials^4^.

This variability in response to clopidogrel is, at least in part, heritable, with genetic factors accounting for up to 70% of the observed variability^5,6^. Inadequate clopidogrel response (also known as clopidogrel resistance) is associated with an increased risk of recurrence of MACE, such as myocardial infarction, stroke, and even death^7–9^.

The first pharmacogenomics study of clopidogrel response published by Mega. *et al.* showed that carrier of *CYP2C19* loss of function (LOF) variants were associated with significantly decreased clopidogrel response by reducing active metabolite of clopidogrel^10^. The first genome-wide association study (GWAS) of clopidogrel response in an European-ancestry cohort was published by Shuldiner *et al.* and similarly found that LOF alleles in *CYP2C19* (*CYP2C19*2*) significantly decreased clopidogrel activity, which resulted in increased risk of post-PCI thrombotic events^6^. This finding has been validated by other studies^11–13^.

It is important to note that most pharmacogenomic studies of clopidogrel have largely been performed in populations of European ancestry, and it remains unclear whether the findings are generalizable to other populations. Additionally, while *CYP2C19* has been shown to explain a significant portion (∼12%) of the variability in clopidogrel response, heritability studies suggest that up to ∼70% of the variance in clopidogrel response is genetic in nature^6^, indicating that other genetic variants likely play a role. Therefore, there is still a need to identify additional genetic variants that contribute to clopidogrel response to improve the efficacy and safety of clopidogrel treatment.

The African American (AA) population is an admixed population which contains more than one ancestry genome. A study done in 24 USA hospitals showed that African Americans (AAs) have a much higher one-year mortality rate (∼7.2%) compared with European-ancestry (∼3.6%) who were treated with clopidogrel^14^ due to the poor antiplatelet response^15,16^. Current GWAS studies often exclude individuals of AA ancestry,^17^ which can lead to a lack of knowledge regarding population-specific genetic variants that may be associated with clopidogrel response in this population. This knowledge gap may limit our ability to fully understand the genetic factors that influence drug response and to develop personalized treatment strategies for AA patients.

The aim of this study is to identify novel biomarkers of clopidogrel response in AAs through GWAS and transcriptomics analysis followed by *in vitro* functional analysis of implicated variants and genes.

## Materials and Methods

### Study populations and sample collection

Participants were recruited through AA Cardiovascular Pharmacogenomics Consortium (ACCOuNT) as previously described^18^. The study protocol was approved by the University of Chicago, University of Illinois, Chicago, Northwestern University, George Washington University, and the DC Veterans Administration Hospital Institutional Review Boards. All participants provided written consent prior to the start of study procedures. In this study, 154 participants were administered a daily dose of 75 mg of clopidogrel for a minimum of 15 days. Blood samples were collected into sodium citrate-vacutainers and analyzed within 4 hours of collection at Northwestern Memorial Hospital diagnostic laboratory, the Washington, DC Veterans Affairs Medical Center or at George Washington University. Samples were tested using the VerifyNow platform, which provided a P_2_Y_12_ reaction units (PRU) value for each sample. Participants with a PRU value of 230 or higher were classified as having high on-treatment platelet reactivity (HTPR). This well-established definition of HTPR has been used in clinical practice and research to identify individuals who may be at increased risk of MACE^19^. Other blood samples were collected into EDTA-vacutainers for DNA isolation and into PAXgene-vacutainers for RNA isolation.

### Genotyping and imputation

DNA was isolated from blood sample using the Gentra Puregene Bood kit from Qiagen (Maryland, USA). DNA samples were genotyped using the Illumina Infinium Multi-Ethnic Global Kit (MEGA) at the University of Chicago Functional Genomics Core using standard protocols. Genotyping outputs were created by Genome Studio using 0.15 GenCall score cutoff.

Standard quality control (QC) procedures were implemented and included: removing the SNPs with an excessive missingness (> 5%) and individuals with missing genotypes (> 5%) or high heterozygosity rates (three standard deviations (SD) away from the mean rate). Hardy-Weinberg equilibrium (HWE) was calculated for all genotyped SNPs. Any SNPs that deviate from HWE were flagged but not removed. Gender misspecification was checked using X chromosome heterozygosity. Any individuals who did not match known sample data were excluded. Identity-by-descent (IBD) checks were performed to identify sample duplicates, contaminated samples, and cryptic relationships. For each pair of samples with estimated IBD coefficients greater than 0.185, only the sample with the highest call rate were retained. Principal component analysis (PCA) was performed to facilitate verification of ancestry and correction for population substructure including HapMap samples from three populations (CEPH (Caucasians), Asians and Yorubas (Africans)) as references.

After the quality Control measures, genotyping data was imputed using the Trans-Omics of Precision Medicine Program (TOPMed) imputation server (https://imputation.biodatacatalyst.nhlbi.nih.gov/#!), using TOPMed imputation panel^20^. SNPs with minor allele frequencies (MAF) below 0.05 and imputation scores under 0.8 were removed. This resulted in 8,599,814 SNPs used in the GWAS analysis.

### Genome-wide association studies (GWAS) analyses

Multivariable logistic regression GWAS was performed to estimate the odds ratios (ORs) and 95% confidence intervals (CI) of each variant for the risk of HTPR (case/control analysis) adjusting for age, gender, principal component 1 (PC1), and PC2 using SNPTEST v2 software. Additionally, multivariable linear regression GWAS was performed to identify the association of SNPs and the change of PRU value as a continuous phenotype.

We also conducted local ancestry inference (LAI) GWAS using Tractor. Briefly, local ancestry at an individual level were estimated used RFMix v2^21^ with a window size of 0.2cM, number of generations equal to 8 and the number of trees to generate per Random Forest set to 100. 1000 Genome African ancestry (YRI: Yoruba in Ibadan, Nigeria), and European ancestry (CEU: Northern Europeans from Utah) populations were used as reference panel. The RFMix v2 ancestry calls were converted to Tractor format, which includes genotype dosage and haplotype count information for African, and European ancestry at each SNP position. The Tractor local ancestry GWAS was then performed using PLINK. This analysis allowed us to use the LAI at each SNPs position as a SNP-based covariate in the GWAS for this admixed cohort. All clinical covariates that showed association to PRU or HTPR were included as covariates in the analysis. The deconvoluted model within TRACTOR was used in this analysis^22^. Meta-analyses on the deconvoluted AFR, and EUR summary statistics was conducted using METAL using a random-effect model.

SNPs with p-value ≤ 5×10^−8^ were considered as genome-wide significance, whereas SNPs with p-value ≤ 5×10^−6^ were considered as suggestive level of significance.

### Validation in Million Veterans Program Clopidogrel cohort

To validate our genome-wide suggestive hit, we used data collected from the Million Veteran’s Program (MVP)^23^ for a cohort of African American patients treated with clopidogrel after a PCI performed at VA catheterization facilities (n=823). Compared to the ACCOuNT cohort, the only clinically relevant phenotype to clopidogrel response available was the Major Adverse Cardiovascular Events (MACE) composite phenotype defined as all-cause death, recurrent MI, and stroke following the index PCI, which was collected from electronic medical records data as previously described.^24^ We extracted genotypes from the MVP Release 4 imputed genomic data; quality of African ancestry imputation in this version of the MVP dataset was improved using the African Genomic Resources (AGR) reference panel.^25^ The frequencies of the alleles and the imputation quality for the genomic hits tested are provided in Table Supplement 2. We tested the association of the carrier status of candidate hits (coded as an indicator for carrying two copies of the effect allele versus 1 or 0 copy as the reference) using a Cox proportional hazard model adjusted for age at procedure, type of stent received (bare metal versus drug eluting), presence of chronic kidney disease (yes/no), prior history of PCI (yes/no) and timing of PCI relative to enrollment in the MVP study (before versus after). As a sensitivity analysis we tested the inclusion of an indicator for PCI indication (acute coronary syndrome, or ACS, versus stable ischemic heart disease) in the model. The effect of ACS was found not significant in the model, and we did not retain the variable in the final model. Statistical analyses were performed in R using the packages ‘survival’ and ‘survminer’^24^.

### HUVEC cell culture, sample collection, and analysis of SNP association to *THSD7A* gene

HUVEC cell lines from one white donor and seven African American (AA) donors were purchased from several vendors: one white and 5 AA from Lonza (Walkersville, MD, USA); 1 AA from ACGT (Manassas, VA, USA); and 1 AA from PromoCell (Heidelberg, Germany).

All HUVEC cell lines were cultured in EGM-Plus medium with supplements (Lonza Walkersville) at 37° C in a 5% CO_2_/95% air cell culture incubator.

DNA was isolated from these 8 cell lines and sequenced to identify their rs7792678 and rs7811849 genotypes. DNA isolation was done using the PureLink™ Genomic DNA Mini Kit (ThermoFisher). Then, DNA was amplified using the Platinum™ SuperFi II Green PCR Master Mix (ThermoFisher) and the PCR product sequenced by ACGT Inc. (Wheeling, IL). The primers used for both PCR amplification and sequencing of *THSD7A* were: tgtgtcaaaacccaaaaggc (forward) and tcccaccatcaactttgggta (reverse).

Total protein and total RNA were isolated from cells cultured in 10-cm plates. The cells from half of one plate were used for protein extraction, and the cells from the other half of the same plate were used for total RNA extraction.

### Differential gene expression analysis of *THSD7A* in GEO data

To investigate the association of expression of *THSD7A* gene and risk of HTPR, the GSE32226^26^ gene expression dataset was downloaded from NCBI GEO (www.ncbi.nlm.nih.gov/geo).^27^ The GSE32226 dataset included exon array data in peripheral blood cells from 26 CAD patients treated with clopidogrel (75 mg/day). All samples were normalized by robust multiarray average (RMA) procedure. For this study, we selected PRU value ≥ 230 as clopidogrel non-response group, while those PRU value ≤ 230 as clopidogrel response group (as defined in our study). The GEO2R interactive web tool was used for difference gene expression analysis. The raw CEL data was analysis by limma^28^ (Linear Models for Microarray Analysis). Only *THSD7A* gene expression data was extracted.

### RNA-sequencing and Quality control analysis (QC)

Blood total RNA was isolated using the PAXgene Blood RNA kit from PreAnalytiX (Qiagen). RNA samples were then depleted of globin mRNA with the GLOBINclear kit (Invitrogen, Thermo Fisher) and RNA quality measured by Bioanalyzer or TapeStation (Agilent, Santa Clara, CA).

Samples with RNA integrity number (RIN) score > 7 were selected for sequencing. TruSeq Stranded mRNA Library Prep Kits, and TruSeq RNA Indexes plate for 96 samples (Illumina, San Diego, CA) were used to prepare libraries for sequencing. The libraries were sequenced using the NovaSeq (Illumina) platform by Novogene (Sacramento, CA) following the manufacturer’s instructions, generating paired-end 50 bp reads with approximately 50 million reads per sample.

Sequencing data quality was assessed using FastQC. The sequencing reads were trimmed to remove adaptor-related sequences and reads with ≥ 3 consecutive bases of aggregate quality score < 20 using Trimmomatic (version 0.39)^29^. The high-quality reads were then quantified at transcript level against the human reference transcriptome (Homo_sapiens.GRCh38.cdna.all.fa) using Salmon^30^.

### Differential gene expression analysis

The differential gene expression (DGE) analysis was performed using R package “DESeq2”^31^. Gene expression values were filtered based on expression thresholds of at least 10 reads in at least 20% of samples. Only protein coding genes and lncRNAs were included in the analysis. The expression values for each gene were normalized using regularized log transformation. Surrogate variable analysis (SVA) was used to account for unmeasured covariates, and all relevant surrogate variables were added to DESeq2 and generalized linear regressions were fit for each gene^32^. Genes with a fold change greater than 2.5 and false discovery rate (FDR) less than 0.05 between HTPR and non-HTPR groups were considered differentially expressed.

### shRNA knockdown of *LAIR1* in MEG-01 cells

The MEG-01 cells were cultured in RPMI 1640 with 10% fetal bovine serum (FBS) and 2 mM l-glutamine plus 100 U/mL penicillin and 100 μg/mL streptomycin in a 37°C humidified atmosphere containing 5% CO_2_/95% air. *LAIR1* shRNA lentivirus clones were purchased from the RNAi Consortium (Sigma-Aldrich). MEG-01 cells were infected by LAIR1 shRNA lentivirus clones. The pLKO.1-puro non-Mammalian control transduction particle (SHC002V) was used as negative control. Puromycin (2ug/ml) was used to select the infected cells. After selection with puromycin (2 ug/ml) for 4 days, real-time quantitative polymerase chain reaction (RT-qPCR) was performed to test the knockdown efficiency.

### RT-qPCR analysis

Total RNA was isolated from MEG-01 cells and HUVECs using the miRNeasy Mini kit (Qiagen, Valencia, CA). Then, RT-qPCR analysis was performed using the Power SYBR® Green RNA-to-CT^TM^ 1-Step Kit (Thermo Fisher, Waltham, MA, USA), in an Applied Biosystems QuantStudio 7 system (Thermo Fisher). PCR primers for human genes *LAIR1* (leukocyte associated immunoglobulin like receptor 1), *SRC* (SRC proto-oncogene, non-receptor tyrosine kinase), *SYK* (spleen associated tyrosine kinase), and *AKT1* (AKT serine/threonine kinase 1) were purchased from Sigma-Aldrich (Saint Louis, MO, USA). The primers sequence for each gene is shown in **Table** S**1**. PCR primers for the human *THSD7A* gene were predesigned (Bio Rad, qHsaCID0007366).

Relative gene expression was quantified using the comparative Ct method (2^−ΔΔCT^), with glyceraldehyde 3-phosphate de-hydrogenase (GAPDH) used as an internal control.

### Western blotting and ELISA analyses

Total protein was extracted using a routine procedure. Briefly, total protein from non-*LAIR1* knockdown and *LAIR1* knockdown MEG-01 cells was extracted using RIPA buffer containing protease and phosphatase inhibitors. Total protein from HUVECs was extracted using RIPA buffer with protease inhibitors. Protein concentration was determined using a Bradford protein assay kit (ThermoFisher Scientific). Lysates were loaded into NuPAGE™ 4–12%, 1.5 mm, 10 well Mini Protein Bis-Tris gels (ThermoFisher Scientific; NP0335BOX), and run at 100 V for 90 min. Gels were then wet transferred to Immun-Blot® PVDF Membrane (Bio-Rad, Hercules, CA) using NuPAGE™ transfer buffer (ThermoFisher Scientific; NP00061) with 10% methanol for 1 hour and blocked for 1 hour at room temperature in Blotting-Grade Blocker Buffer (Bio-Rad, Hercules, CA). Primary antibodies were diluted with their respective blocking buffers and incubated overnight at 4 °C. Washes were performed with TBS 0.1% Tween-20 (TBST) before the addition of secondary antibody for one hour at room temperature. Washes were performed with 1× TBST before imaging on Odyssey Fc. Protein detection was performed using Image Studio Ver 5.2. Most primary antibodies were purchased from Santa Cruz Biotechnology (Dallas, TX), including LAIR-1, Src, Phospho-Src (Tyr419), ERK ½, Phospho-ERK ½, and Cell Signaling (Danvers, MA), including phospho-Akt (Ser473), Akt, phospho-Syk (Tyr525/526), Syk, and GAPDH. THSD7A protein expression in HUVECs was also confirmed by ELISA (MyBioSource, San Diego, CA).

All values were represented as mean ± standard deviation (SD) from at least three independent experiments. Statistical differences between groups were measured by using Student’s T-test. P-values in all experiments were considered significant at less than or equal to 0.05.

## Results

The overview of the study can be seen in Figure 1. This study included 154 subjects from the ACCOuNT cohort with genotype data, of which 99 had corresponding RNA-seq data. After QC for genotyping and RNA-seq, 141 samples passed QC for genotype data (case 38, Controls 103 – Table 2). Among 99 RNA-seq data, 93 included PRU value (22 cases and 71 controls). In addition, we validated our finding in a cohort from 823 African American MVP participants on clopidogrel after a PCI. The characteristics of both patient cohorts are summarized in the **Table 1**.

**Figure 1:**
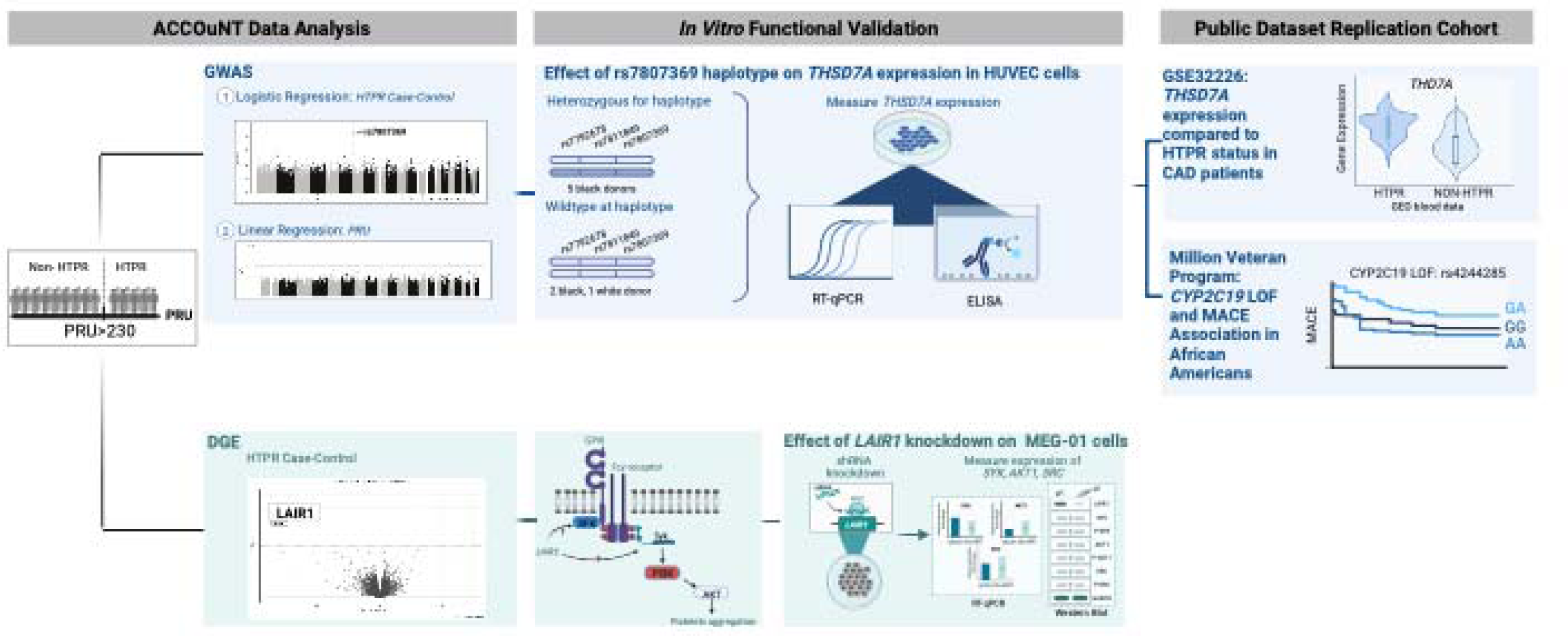
Overview of Study. Infographic showing the multiple stages of this study including discovery cohort analysis, functional *in vitro* follow up and validation cohorts for both the genomic (blue) and transcriptomic (green) findings.

**Table 1.**
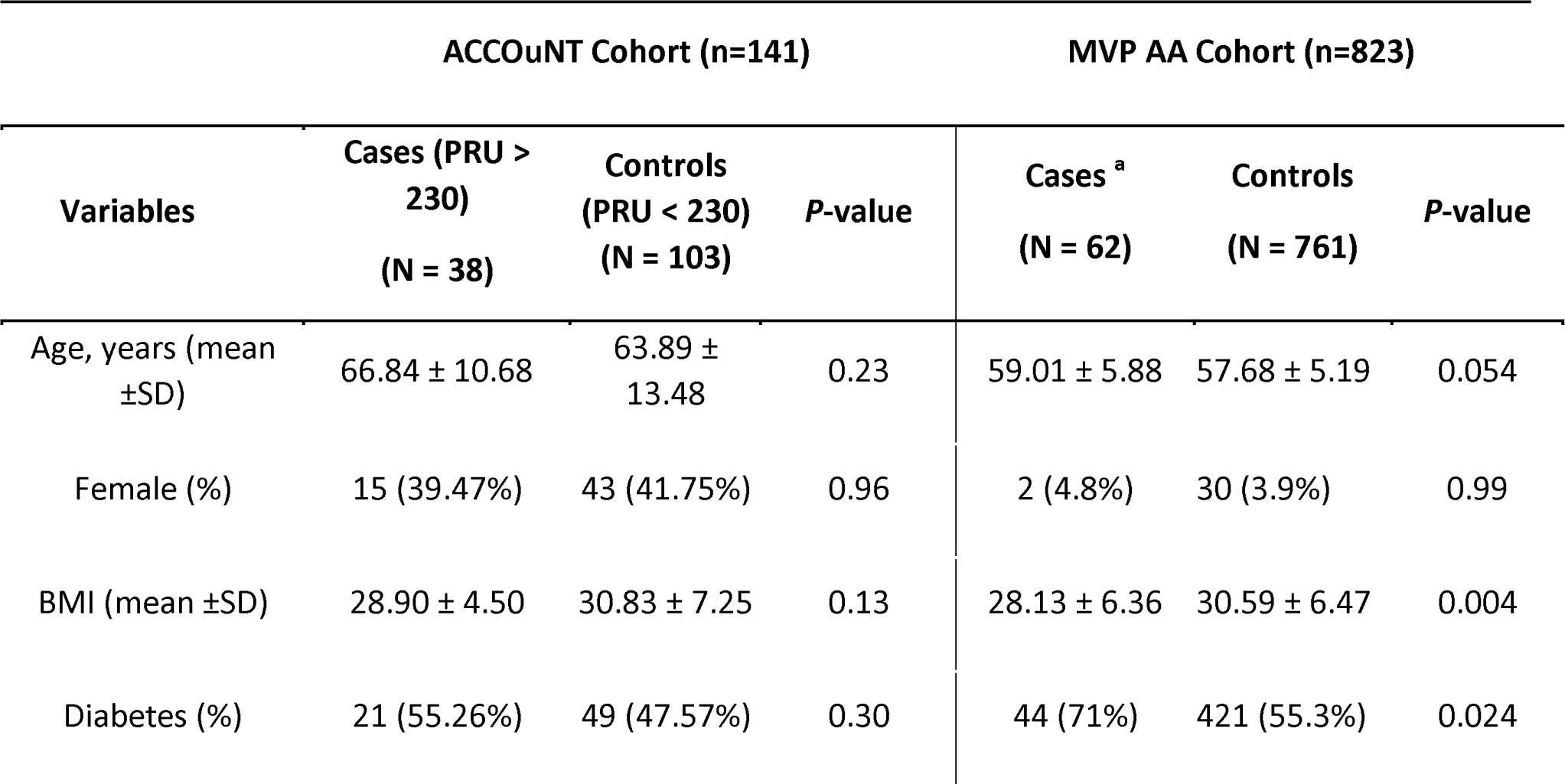
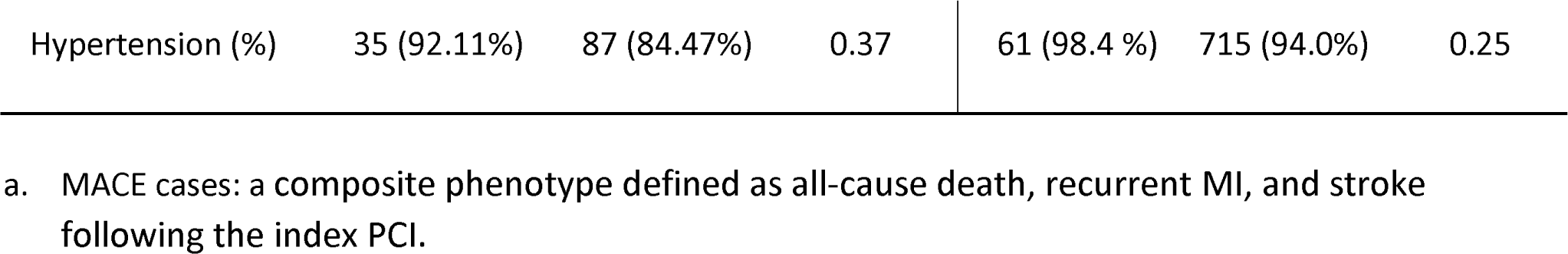
Demographic table.

| ACCOuNT Cohort (n=141) |  |  |  | MVP AA Cohort (n=823) |  |  |
| --- | --- | --- | --- | --- | --- | --- |
| Variables | Cases (PRU > 230)<br>(N = 38) | Controls (PRU < 230)<br>(N = 103) | P-value | Cases <sup>a</sup><br>(N = 62) | Controls<br>(N = 761) | P-value |
| Age, years (mean ±SD) | 66.84 ± 10.68 | 63.89 ± 13.48 | 0.23 | 59.01 ± 5.88 | 57.68 ± 5.19 | 0.054 |
| Female (%) | 15 (39.47%) | 43 (41.75%) | 0.96 | 2 (4.8%) | 30 (3.9%) | 0.99 |
| BMI (mean ±SD) | 28.90 ± 4.50 | 30.83 ± 7.25 | 0.13 | 28.13 ± 6.36 | 30.59 ± 6.47 | 0.004 |
| Diabetes (%) | 21 (55.26%) | 49 (47.57%) | 0.30 | 44 (71%) | 421 (55.3%) | 0.024 |
| Hypertension (%) | 35 (92.11%) | 87 (84.47%) | 0.37 | 61 (98.4 %) | 715 (94.0%) | 0.25 |

**Table 2.** Top SNPs of GWAS analysis of HTPR.

| HTPR |  |  |  |  |  |  |  |  |  |
| --- | --- | --- | --- | --- | --- | --- | --- | --- | --- |
| SNPID | Chr | Position | Non-effect allele | Effect allele | MAF | OR | 95% CI | SE | Pval |
| rs7807369 | 7 | 11465339 | C | G | 0.31 | 4.02 | 2.30 - 7.01 | 0.46 | 4.56E-09 |
| rs11983224 | 7 | 11465666 | T | C | 0.32 | 3.65 | 2.10 - 6.33 | 0.43 | 5.11E-08 |
| rs10224183 | 7 | 11460107 | C | T | 0.36 | 3.75 | 2.17 - 6.50 | 0.39 | 7.69E-08 |
| rs6013452 | 20 | 52407102 | A | T | 0.46 | 3.53 | 2.02 - 6.17 | 0.40 | 2.65E-07 |
| rs58975724 | 16 | 8656828 | A | G | 0.24 | 3.84 | 2.15 - 6.88 | 0.42 | 3.09E-07 |
| rs7793779 | 7 | 11459627 | G | A | 0.33 | 3.37 | 1.95 - 5.84 | 0.41 | 3.16E-07 |
| rs10094297 | 8 | 18524850 | T | C | 0.36 | 0.21 | 0.11 - 0.43 | 0.40 | 3.64E-07 |
| rs10224154 | 7 | 11459952 | G | A | 0.32 | 3.29 | 1.90 - 5.71 | 0.42 | 4.08E-07 |
| rs58075729 | 4 | 26841202 | C | A | 0.25 | 3.82 | 2.15 - 6.81 | 0.40 | 5.41E-07 |
| rs4845158 | 1 | 190008728 | G | A | 0.50 | 0.29 | 0.17 - 0.52 | 0.36 | 9.74E-07 |
| rs7792678 | 7 | 11459151 | A | C | 0.36 | 3.16 | 1.84 - 5.45 | 0.41 | 1.11E-06 |
| rs7811849 | 7 | 11459290 | T | A | 0.36 | 3.00 | 1.74 - 5.16 | 0.41 | 2.67E-06 |
| rs1149515 | 7 | 16736755 | T | C | 0.14 | 15.61 | 2.36 - 103.04 | 1.05 | 2.74E-06 |

Chr: chromosome; MAF: minor allele frequencies; OR: odds ratio; 95% CI: 95% confidence interval; SE: standard error

### GWAS identified association at the *THSD7A* gene locus

Our logistic model GWAS in AA patients treated with clopidogrel identified one locus that attained genome-wide significance level (p<5* 10^−8^). The top SNP rs7807369 on chromosome 7 was associated with an increased risk of HTPR (OR: 9.11, 95% confidence interval (CI): 3.66 – 22.67, p = 5.57 * 10^−9^) (**Figure 2, Table 2**). Our GWAS association to PRU (as a continuous phenotype) did not identify any genome-wide significant signals. One signal located on chromosome 18 reached the suggestive significance level (p<1×10^−6^). The top SNP rs2226859 was associated with decreasing the PRU value (**β** = −0.61, 95%CI: −0.72 - −0.49, P = 8.41*10^−7^). This SNP is in an intergenic region and is not eQTL to any gene in the GTEx database. Our LAI GWAS of HTPR did not find any genome-wide significant hits. However, 10 SNPs (in 5 loci) reached nominal significance. (**Table 3**)

**Figure 2.**
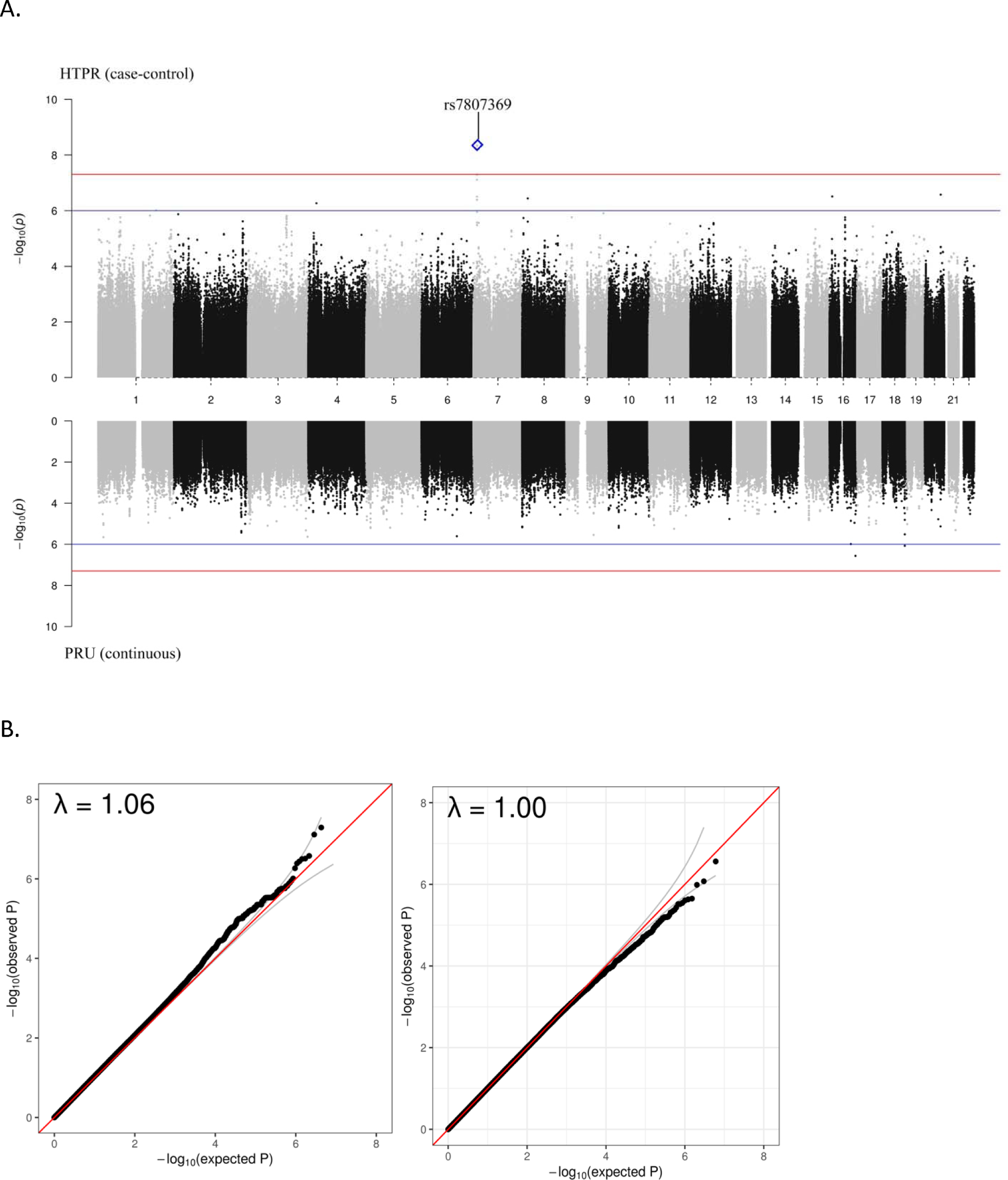
Miami and QQ plots from GWAS. Miami plot representing the association between single nucleotide polymorphisms (SNPs) and High on Treatment Platelet Reactivity (HTPR) (Top) and the association between SNPs and the change of PRU value (Bottom). The x-axis shows the position of each SNP on the chromosome, while the y-axis displays the association (-log_10_ P-value) between the SNP and the trait of interest. The red line represents the threshold for genome-wide significance (p = 5*10^−8^), and the blue line represents the suggestive level of significance (p = 1*10^−6^). The QQ plot displays the observed p-values of the genome-wide association study (GWAS) summary data on the y-axis, compared to the expected p-values under the null hypothesis of no association on the x-axis. Deviations from the diagonal line indicate departures from the null hypothesis, where points above the line suggest more significant associations than expected, while points below the line suggest fewer significant associations than expected. (A) The GWAS identified one locus (*THSD7A* on chromosome 7) to be associated with higher risk of HTPR at genome-wide significance level (p < 5* 10^−8^). (B) QQ plot for GWAS. The genomic control value for the GWAS was 1.06 for logistic regression model GWAS (Left) and 1.00 for linear regression model GWAS.

**Table 3:** Top SNPs of LAI GWAS analysis of HTPR.

| SNP ID | Chr | Position | Non-effect allele | Effect allele | EAF | OR | 95% CI | SE | P-value |
| --- | --- | --- | --- | --- | --- | --- | --- | --- | --- |
| rs6577698 | 8 | 137007398 | T | C | 0.27 | -1.64 | -2.31 – -4.19 | 0.34 | 1.55E-06 |
| rs11723866 | 4 | 136777689 | A | G | 0.37 | 2.24 | 1.30 – 3.02 | 0.48 | 2.82E-06 |
| rs194873 | 6 | 82453531 | T | G | 0.75 | 1.81 | 1.05 – 2.44 | 0.39 | 2.97E-06 |
| rs977413 | 21 | 24008267 | A | G | 0.41 | 1.74 | 1.01 – 2.35 | 0.38 | 3.53E-06 |
| rs2137705 | 21 | 24013695 | T | C | 0.54 | 1.58 | 0.91 – 2.13 | 0.34 | 3.60E-06 |
| rs1830229 | 21 | 24010134 | T | C | 0.60 | -1.75 | -2.49 – -4.50 | 0.38 | 3.77E-06 |
| rs11721336 | 4 | 136782208 | A | G | 0.63 | -2.21 | -3.15 – -5.62 | 0.48 | 4.49E-06 |
| rs1480155 | 5 | 146431164 | A | G | 0.22 | -2.18 | -3.11 – -5.18 | 0.48 | 4.52E-06 |
| rs6535124 | 4 | 136808619 | A | G | 0.62 | -2.00 | -2.87 – -5.18 | 0.44 | 5.18E-06 |
| rs191786 | 6 | 82497480 | T | C | 0.72 | 1.85 | 1.05 – 2.46 | 0.41 | 6.20E-06 |

Chr: chromosome; EAF: effect allele frequencies; OR: odds ratio; 95% CI: 95% confidence interval; SE: standard error

The regional plot shows that the top SNP and variants in high linkage disequilibrium (LD) from the logistic regression analysis are located at the intronic region of *THSD7A* gene. Based on the output of UCSC Genome browser, two SNPs are in high LD (r^2^ ≥ 0.8) with rs7807369 and are located within a potential enhancer/promoter region (**Figure 3**). Hi-C interactions analysis from Vanno Portal corroborated these findings with SNPs rs7792678 and rs7811849 affect the *THSD7A* expression in Human umbilical vein endothelial (HUVEC) cell line by regulating the activation of the promoter (**Supplement Figure 1**). To investigate the association of *THSD7A* gene expression to the risk of HTPR, we compared the difference in *THSD7A* gene expression using public gene expression data (GSE32226)^26^ comparing HTPR and non-HTPR groups (as defined in our GWAS cohort). From this analysis, higher expression of *THSD7A* was associated with higher risk of HTPR (p = 0.004) (**Figure 4A**).

**Figure 3.**
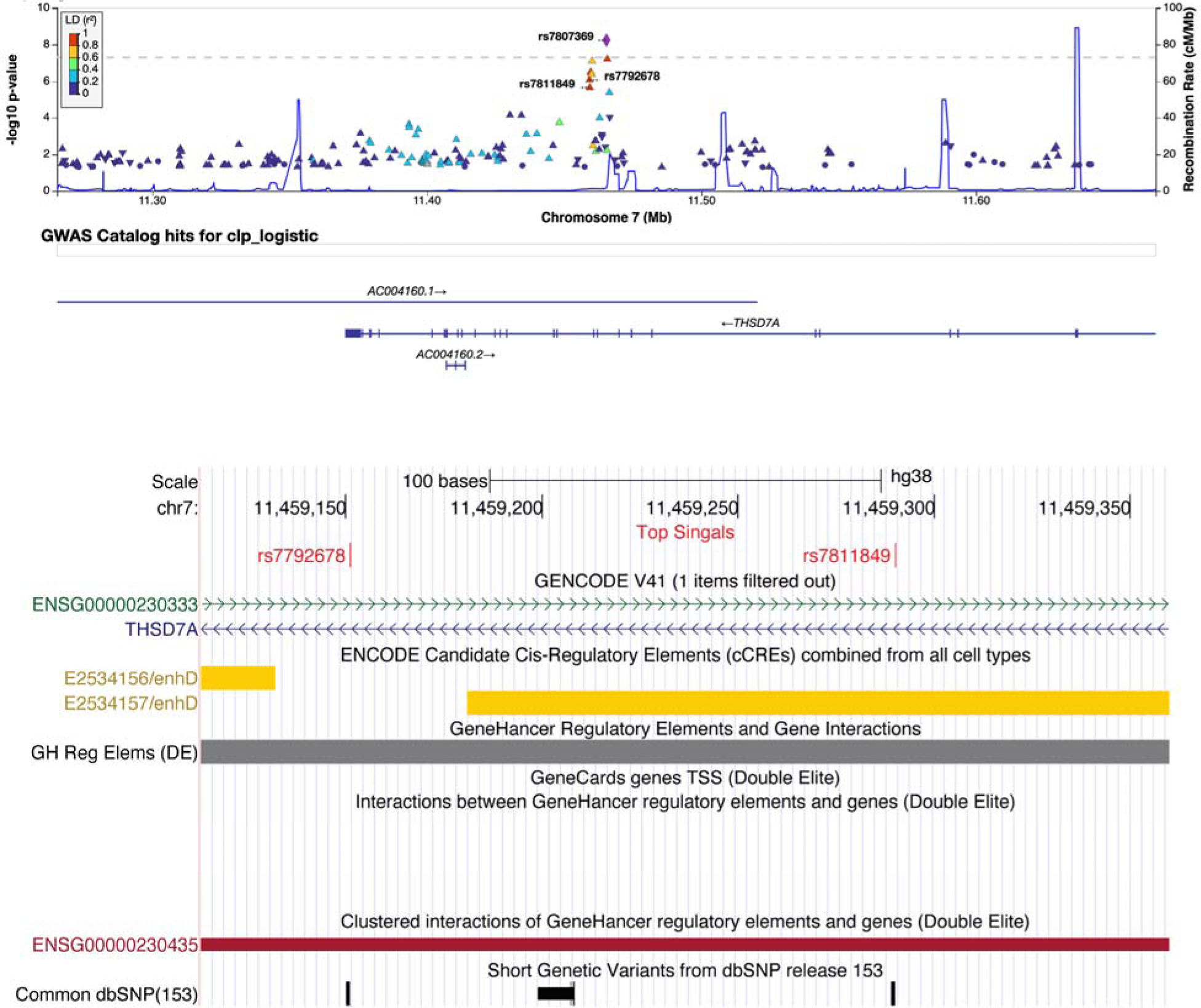
Regional plot of top SNP. The colors of the circles refer to linkage disequilibrium (LD) (r^2^) between top SNP rs7807369 (purple diamond) and nearby SNPs (based on pairwise r^2^ values from the 1000 Genomes Project reference panel). The blue line and right y-axis show the estimated recombination rate. The x-axis represents the genomic position in chromosome 7 and the left y-axis represents the -log10 p of association with HTPR in discovery cohort. Region plot showed *THSD7A* variant rs7792678 is high LD with top SNP rs7807369. Both are located at the intronic region of *THSD7A* gene. SNPs rs7792678 and rs7811849 located at the potential enhancer/promoter region based on ENCODE annotation displayed in the UCSC Genome Browser dataset.

**Figure 4.**
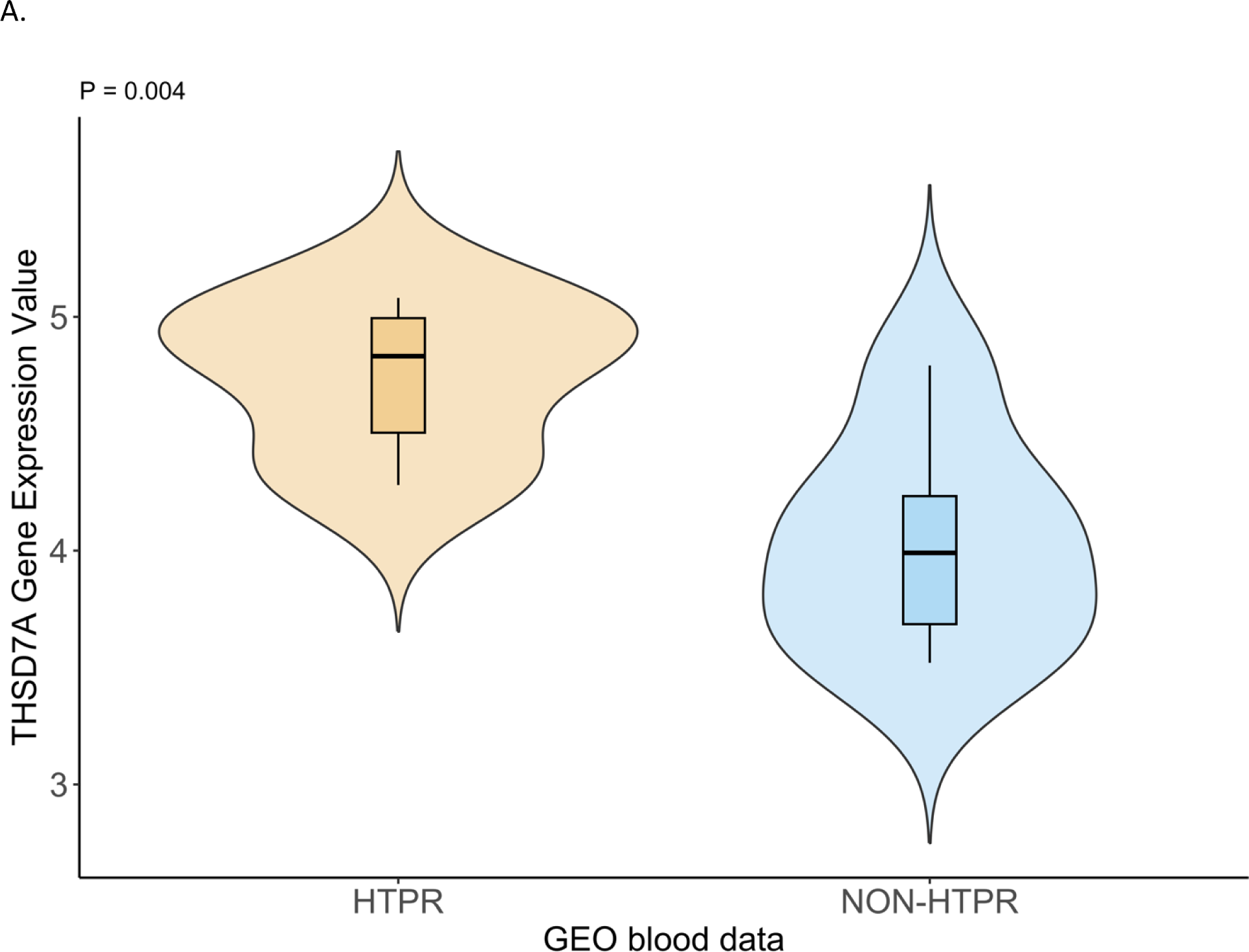
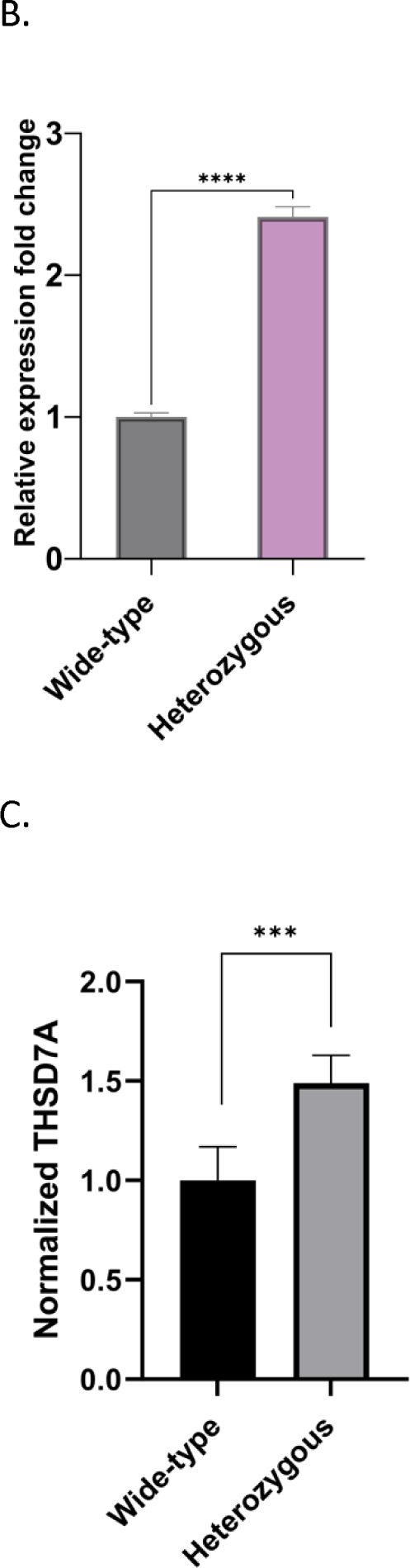
THSD7A expression associated with higher risk of HTPR. (A) The violin box plot of the expression of *THSD7A* gene in HTPR (PRU >230) and non-HTPR groups from the GEO dataset GSE32226. The *THSD7A* gene expression is significant higher in HTPR group compared with non-HTPR group (p = 0.004). (B) Fold-change in expression of *THSD7A* gene in carriers of the effect alleles compared with carriers wild-type alleles in HUVEC cell cultures. Bar heights indicate the mean expression of *THSD7A* gene in groups of white wild-type, AA wild-type, and AA effect alleles carriers. Error bars indicate 95% confidence interval estimates of the mean expressions. (*) = p<0.05, (**) = p<0.01, (***) = p<0.001, (****) = p<0.0001, ns = non-significant. (C) Quantified difference in protein expression from the THSD7A ELISA show in the bar chart for the same groups.

To investigate the potential function of rs7792678 and rs7811849 on *THSD7A* gene expression, we sequenced seven HUVEC (primary endothelial) cell cultures from AA donors and one from a European Ancestry donor. The European and 2 of the AA HUVEC cultures were wild type for the associated haplotype, while 5 AA HUVEC cultures were heterozygous for the associated haplotype. RT-qPCR found that the cultures that carried the effect haplotype showed increased expression of *THSD7A* (2.2-fold compared with the wild type, p = 0.0006) (**Figure 4B**). Using ELISA, we showed that THSD7A protein levels significantly increased by a 50% (p = 0.004) in the cultures carrying the effect haplotype as compared with the protein level in the cells with the wild-type haplotype (**Figure 4C**).

### External Validation of top association in the MVP cohort

We examined the association of the top hits with the occurrence of MACE in a sample of AA MVP participants receiving clopidogrel after a PCI. We restricted this analysis to 823 AA participants under the age of 65 at time of the PCI due to loss of outcomes data in older individuals who are more likely to receive care outside of the VA healthcare system. Distribution of genotypes and MACE events are presented in Table S3. We performed a time-to-event analysis for the occurrence of MACE using a Cox proportional hazard model adjusted for age at procedure, type of stent used, existence of chronic kidney disease, prior history of PCI and timing of PCI and enrollment in the MVP study. We were able validate rs191786 (found in the LAI GWAS of HTPR, Table 3, OR = 1.85, p = 6.21×10^−6^), with a significantly elevated risk of MACE for the homozygous carriers of the C allele in the MVP AA cohort (OR = 1.77, p = 0.049; Table 4). No significant association was found for rs11983224. Homozygous carriers of the C allele for rs1149515 were found at significantly lower risk of MACE (OR=0.54, p=0.0179). Of note the association of *CYP2C19* loss-of-function with MACE (p=0.738) was not replicated in this sample of African American patients in contrast to many of the European studies to date (Table 4)

**Table 4:** Validation of Top SNPs in the MVP cohort using an adjusted Cox model for the risk of Major Adverse Cardiac Event (MACE) among African American patients receiving clopidogrel after a percutaneous coronary intervention (PCI).

| SNPID | Chr | Position | Non-effect allele | Effect allele | EAf | Hazard Ratio | 95% CI | Pval |
| --- | --- | --- | --- | --- | --- | --- | --- | --- |
| **rs7807369** | 7 | 11465339 | C | G | 0.344 | 1.41 | 0.73-2.71 | 0.308 |
| **rs11983224** | 7 | 11465666 | T | C | 0.363 | 1.53 | 0.83 – 2.82 | 0.177 |
| **rs7792678** | 7 | 11459151 | A | C | 0.386 | 1.33 | 0.72-2.46 | 0.361 |
| **rs7811849** | 7 | 16736755 | T | A | 0.382 | 1.37 | 0.74-2.53 | 0.316 |
| **rs1149515** | 7 | 16736755 | T | C | 0.835 | 0.55 | 0.33 – 0.90 | 0.0179 |
| **rs191786<sup>a</sup>** | 6 | 82497480 | T | C | 0.777 | 1.77 | 1.002 – 3.14 | 0.0490 |
| **CYP2C19<sup>b</sup> LOF** | 9 |  |  |  |  | 0.91 | 0.53 – 1.57 | 0.738 |

N=823 patients. EAF, effect allele frequency observed in the sample of African ancestry. Hazard ratios with 95% confidence intervals and P values for the risk of MACE among patients carrying two copies of the effect alleles versus 1 or 0 copy (reference) using a Cox proportional hazard model adjusted for age at procedure, type of stent received (bare metal versus drug eluting), presence of chronic kidney disease (yes/no), prior history of PCI (yes/no) and timing of PCI relative to enrollment in the MVP study (before versus after).
a - Found in the LAI GWAS on HTPR
b – Patients carrying 1 or 2 copies of a loss-of-function *CYP2C19* variant (\*2, \*3, \*4, \*5, \*8)

### Low expression of *LAIR1* was associated with an increased risk of HTPR

The differential gene expression analysis using RNA-seq data from blood samples identified *LAIR1* as associated with HTPR. Lower expression of *LAIR1* was associated with an increased risk of HTPR (**Figure 5**). To assess the role of *LAIR1* in clopidogrel response, shRNA knockdown of LAIR1 by lentivirus shRNA clones was performed in MEG−01 cell (megakaryocyte cell line). The expression of *LAIR1* was significantly reduced with *LAIR1* knockdown. The knockdown of *LAIR1* significantly increased the expression of *SYK*, *AKT1* but not *SRC*. (**Figure 6**).

**Figure 5.**
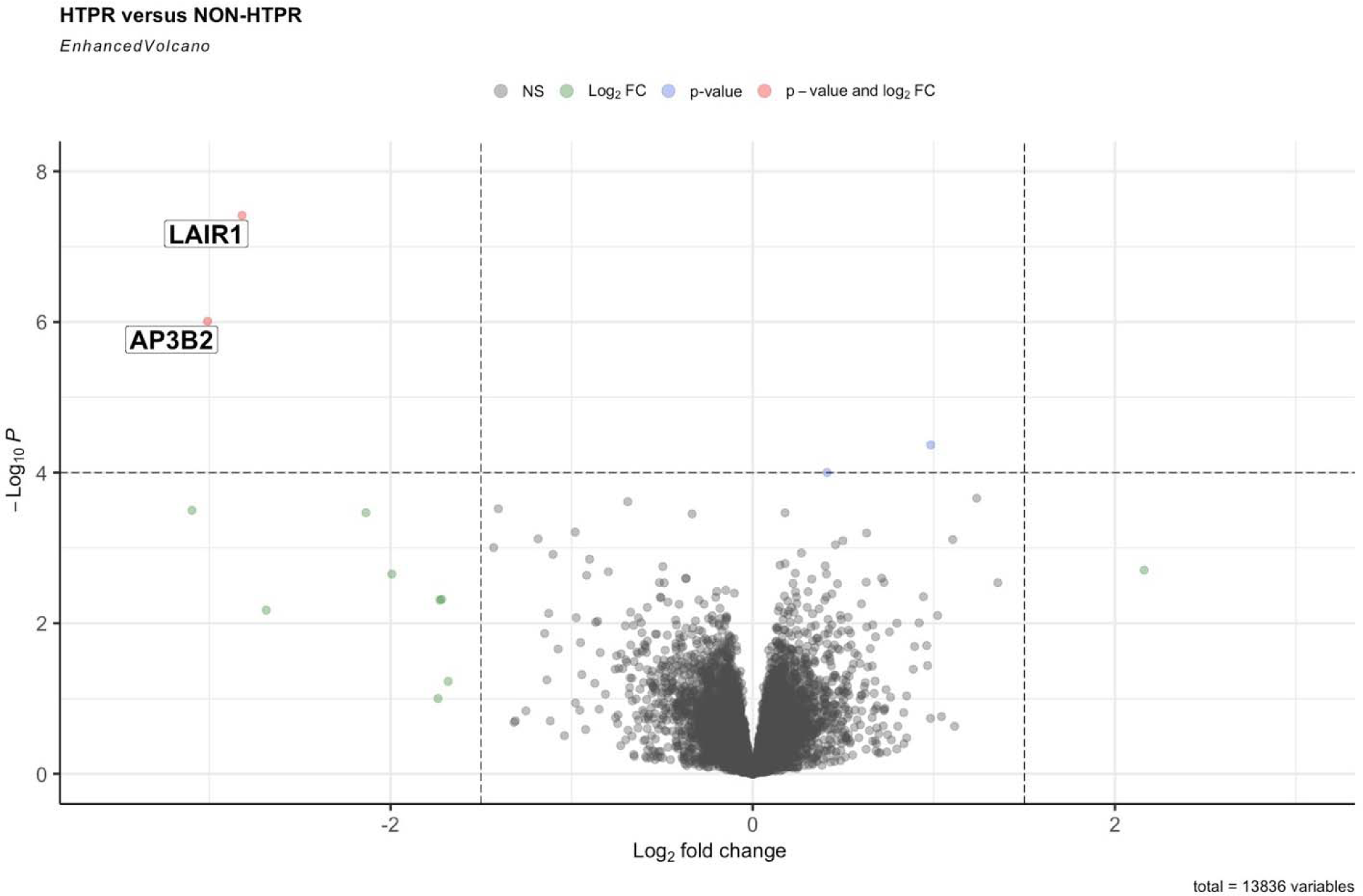
Volcano plot of the differential gene expression analysis. The volcano plot illustrates the statistical significance and fold change for each gene in the RNA-seq dataset from whole blood. Each point represents a gene, with the x-axis representing the log2 fold change and the y-axis representing the negative logarithm of the p-value. The red color point represents the gene with significant p-value (< 4*10^−6^) and log2 fold change.

**Figure 6.**
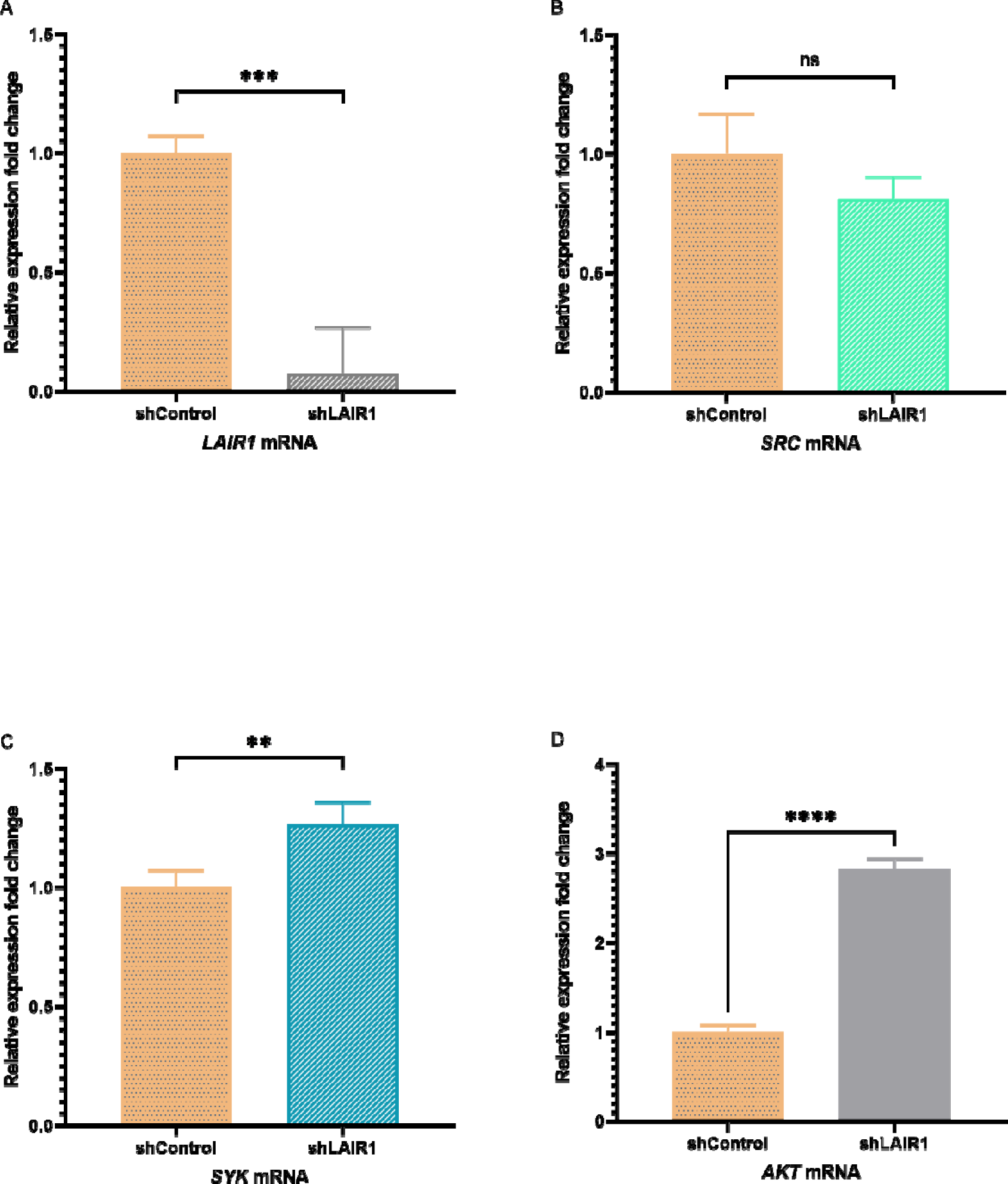
RT-qPCR. Fold-change expression of target genes *LAIR1* (A), *SRC* (B), *SYK* (C), and *AKT* (D), between MEG−01 cells after *LAIR1* knockdown and wild-type cells. Bar heights indicate the mean expression of relative genes. Error bars indicate 95% confidence interval estimates of the mean expressions. (*) = p<0.05, (**) = p<0.01, (***) = p<0.001, (****) = p<0.0001, ns = non-significant. Lack of *LAIR1* expression increased the expression of *SYK* and *AKT* significantly with no change in SRC gene expression.

The effect of *LAIR1* knockdown in protein expression was investigated by Western blot analysis. The results showed that the *LAIR1* knockdown cells exhibited a significant decrease in LAIR1 protein compared with the control group. Moreover, the knockdown of LAIR1 led to an increase in the protein expression levels of Syk and Akt. There is no significant difference for protein level of SRC between LAIR1 knockdown and control groups. Taken together, these data demonstrate that LAIR1 inhibits Syk expression and decreases the downstream AKT pathway (**Figure 7**).

**Figure 7.**
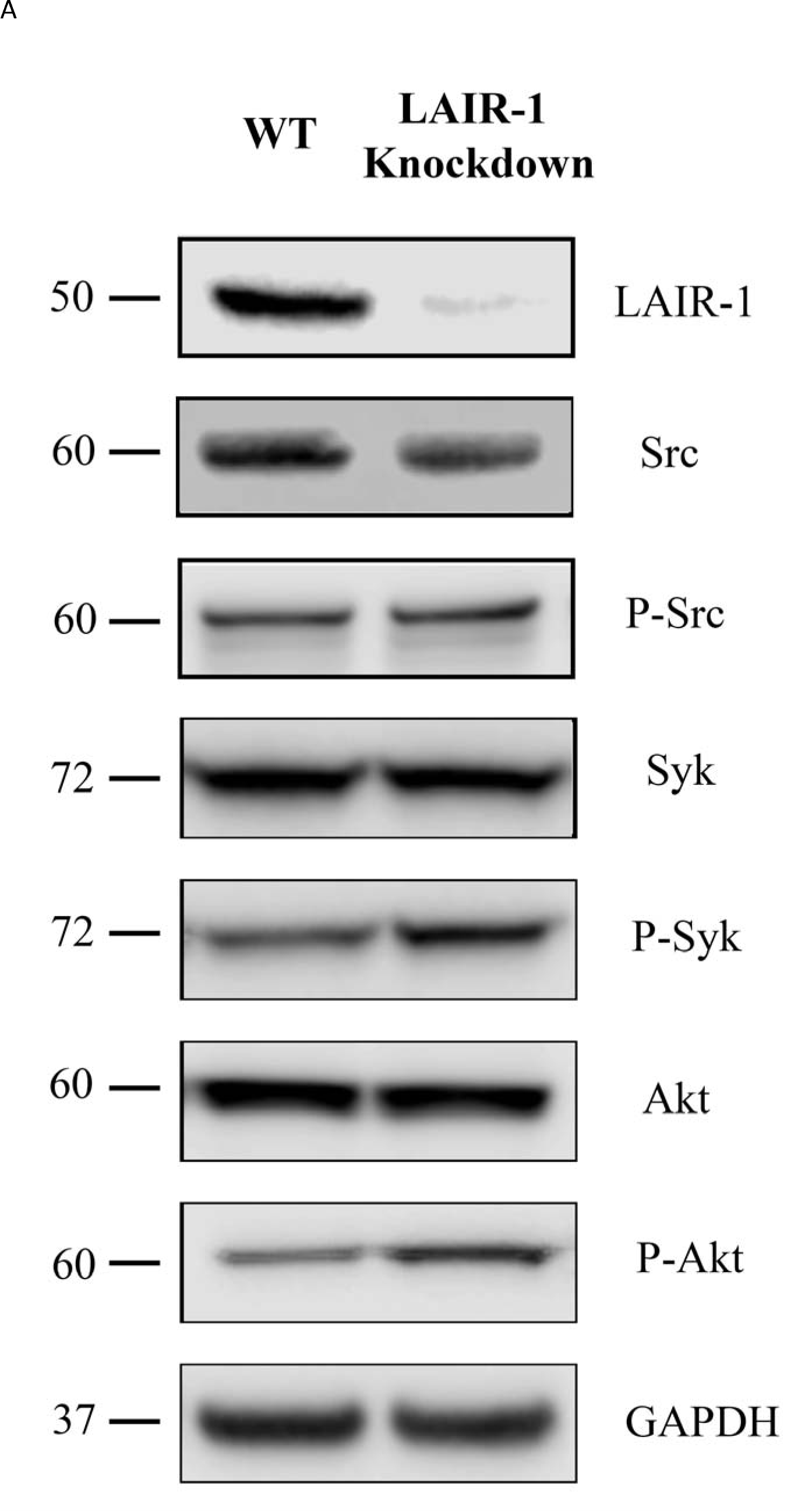
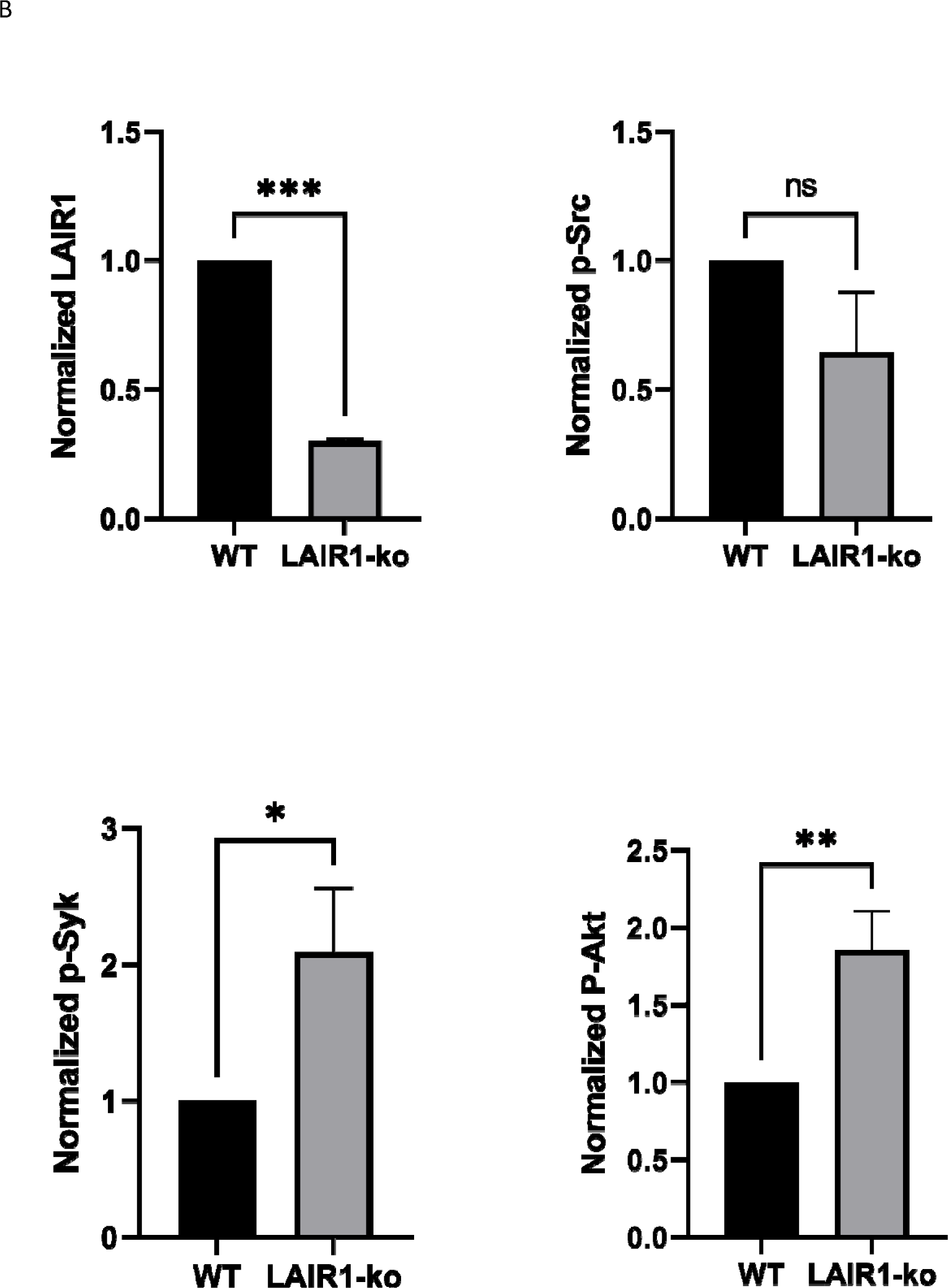
LAIR1 knockdown cells increases AKT pathway. (A) Whole-cell lysates of MEG−01 from wild-type and LAIR1 knockdown groups were Western blotted with anti-LAIR1, anti-SRC, anti-p-SRC, anti-Syk, anti-p-Syk, anti-Akt, anti-p-Akt, and GAPDH antibodies. Representative images are shown. (B) Quantification of western blotting images. Bar heights indicate the mean value of relative proteins. Error bars indicate 95% confidence interval estimates of the mean values. (*) = p<0.05, (**) = p<0.01, (***) = p<0.001, (****) = p<0.0001, ns = non-significant. The LAIR1 knockdown MEG−01 cells show significant lower LAIR1 protein level compared with wild-type MEG−01 cells. The LAIR1 knockdown cells increased the phosphorylation levels of Syk and Akt proteins.

## Discussion

This is the first GWAS study to investigate biomarkers of clopidogrel response in AAs. We identified a potential functional variant, rs7792678, on the potential enhancer/promoter region that is associated with a higher risk of HTPR. According to the gnomAD browser, the frequency of the effect allele for rs7792678 is 0.414 among AA populations, compared to 0.135 among European populations. This may explain the finding of that AAs have a higher clopidogrel resistance compared to Europeans^33,34^. Within the ICPC European clopidogrel study this SNP was found at a frequency of 0.105 and was not associated with PRU.^35^ This study did not investigate genetic associations to HTPR. While we were not able to validate the association of rs7792678 to MACE in the AA MVP cohort, we did find an association to a second SNP (rs1149515) at a nearby locus in Chromosome 7 that did show association to MACE in the AA MVP cohort (**Table 5**). rs1149515 is in moderate LD with an eQTL to the *TSPAN13*, however no known function has been found for this variant.

Using HEUVEC cells derived from AAs, we were able to validate the effect of the C allele of rs7792678 with higher expression of thrombospondin 7A (*THSD7A).* Additionally, the gene expression data from within a multiethnic cohort of clopidogrel patients showed higher expression of *THSD7A* in patient with HTPR as compared with the non-HTPR group. These validation efforts point to a potential role for *THSD7A* in clopidogrel response. *THSD7A* has been implicated in several disease process, including venous thromboembolism^36^, and CAD^37^, both of which are linked to platelet function. *THSD7A* encodes a protein that play a crucial role in endothelial cell migration and angiogenesis^38^. Studies have shown that *THSD7A* can inhibit primary endothelial cell migration and tube formation^39^, which would result in the dysfunction of the endothelial cells. Previous studies have shown endothelial dysfunction was significantly associated with HTPR in patients who were treated with clopidogrel^40–42^.

Notable, we did not detect an association of clopidogrel response with the *CYP2C19*2* allele. *CYP2C9*2* was present in our cohort (MAF = 0.177). However, it showed no association to either HTPR or PRU (P = 0.60 and 0.62). *CYP2C19*2* carriers, a LOF variant of *CYP2C19*, were reported to have a decreasing response to clopidogrel in European and Asian populations^6,43^. While it is clear that the *CYP2C19*2* variant has functional consequences on CYP2C19 enzyme function^44^, our lack of association suggests that additional factors (including additional SNPs) may modify this effect in African ancestry populations. More in-depth sequencing is needed to better understand the role of this clinically actionable SNP on clopidogrel response in African ancestry populations, who carry genetic variation not found in other populations. Moreover, the Cox regression analysis also showed no association of *CYP2C19* LOF alleles to MACE in this independent African American cohort. More in-depth sequencing is needed to better understand the role of this clinically actionable SNPs on clopidogrel response in African ancestry populations.

This study identified *LAIR1* expression is associated with a decreased risk of HTPR. *LAIR1* is a transmembrane glycoprotein that is expressed on hematopoietic and megakaryocytes cells (the precursor cell to platelets)^45,46^. GPVI is a platelet specific collagen receptor that plays a crucial role in platelet activation and aggregation^47^. Both LAIR1 and the gene encoding GPVI (glycoprotein VI) are located on chromosome 19 – making up the leukocyte receptor complex (LRC)^48^. The collagen-binding site on LAIR1 is similar to that of GPVI, a well-known platelet receptor that initiates platelet activation and aggregation^45^. Unlike GPVI, LAIR1 is an inhibitory receptor that binds collagen^49^. A recent study by Smith *et al*.^50^ demonstrated that mice with *LAIR1* deficiency showed a 25% increase in platelet counts, a longer platelet half-life, and increased platelet formation. Our study provides evidence that knockdown of *LAIR1* in human megakaryoblast cells results in increased activation of the GPVI pathway. Specifically, we found that SYK and AKT gene and protein expression levels were significantly increased with *LAIR1*-knockdown. SYK and AKT are two important factors in the platelet aggregation pathway.^51^ These findings suggest that LAIR1 and GPVI may co-regulate platelet aggregation in human cells. Based on the abovementioned evidence, LAIR1 is a promising target gene that regulates platelet aggregation.

### Limitations

One of the major limitations of this study is the small sample size used for analysis. The limited sample size may have reduced the statistical power to detect small to medium effect sizes and rare variants, potentially limiting the generalizability and precision of the findings. We have tried to remedy this limitation through functional validation of our finding in *in vitro* models as well as in independent data sets. The second limitation is the lack of replication study to validate our GWAS result. Replication is unfortunately not possible as no other independent AA clopidogrel dataset exists. The ICPC cohort is a European cohort which showed that most of our significant SNPs were below 5% allele frequency in their cohort and hence not included in the analysis. Of the SNPs that were also tested in this cohort, none were associated to PRU. They may be due to differences in LD structure between Europeans and African populations as well as differences in phenotyping (no HTPR associations were tested in the ICPC cohort). Future studies with larger AA and multi-ancestry cohorts are warranted to validate and expand upon the findings reported here.

In conclusion, we have performed the first GWAS of clopidogrel response in an African American cohort. These data will be useful as we move toward in the implementation of genotypes-guided clopidogrel therapy in diverse clinical populations. Our work shows that the effect of known variants differs by population and potentially by ancestry.

## Data Availability

Genomic and de-identified clinical data for the ACCOuNT participants will be shared on the GEN3 platform at https://gen3.datacommons.io/discovery/ACCOuNT_Clopidogrel_Arm through a control access application. All relevant genetic data for the MVP cohort are available in the main paper and supplemental data. Individual data cannot be shared publicly according to the Data Access Policy of the Million Veteran Program in the VA Office of R&D in Veterans Health Administration.

## Disclaimer

This publication does not represent the views of the Department of Veterans Affairs or the United States Government.

---

**Table S1.**
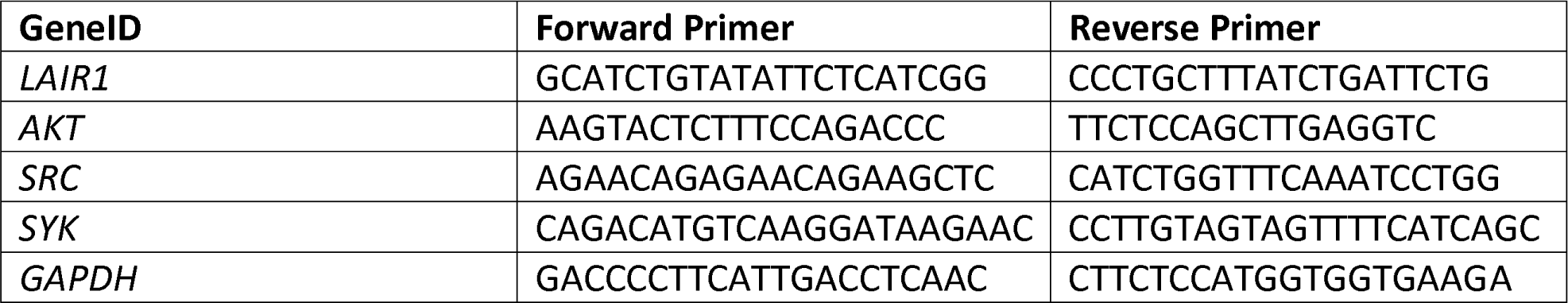
Primers for RT-qPCR.

**Table S2:** Frequencies of alleles and quality of imputation of the Top SNPs in the MVP dataset (Release 4)

| rs ID | Analysis | AF AFR | R<sup>2</sup> AFR | HWE P |
| --- | --- | --- | --- | --- |
| rs7807369 | Log GWAS | 0.35117 | 0.856623 | 0.47614 |
| rs11983224 | Log GWAS | 0.373517 | 0.854156 | 0.346887 |
| rs7792678 | Log GWAS | 0.399141 | 0.921888 | 0.348683 |
| rs7811849 | Log GWAS | 0.394364 | 0.925986 | 0.176193 |
| rs1149515 | Log GWAS | 0.825222 | 0.829126 | 0.594086 |
| rs191786 | LAI GWAS | 0.786334 | 0.901932 | 0.873431 |

The three candidate SNPS were imputed using the African Genome Resources (AGR) reference panel in MVP Release 4.; AF AFR, variant allele frequency among MVP participants of African ancestry; R<sup>2</sup> AFR, quality of imputation among MVP participants of African ancestry; HWE P, P values for the Hardy-Weinberg Equilibrium test (non-significant P values indicate that the equilibrium is respected at that position)

**Table S3:** Distribution of genotypes and MACE rate for the Top SNPs in the MVP sample of African American patients receiving clopidogrel after a PCI.

|  | Overall (n=823) | Cases (n=62) | Controls (N=761) | MACE rate (%) | P value |
| --- | --- | --- | --- | --- | --- |
| **rs7807369** |  |  |  |  |  |
| CC | 359 (43.6) | 26 (41.9) | 333 (43.8) | 7.24% | 0.407 |
| CG | 362 (44.0) | 25 (40.3) | 337 (44.3) | 6.91% |  |
| GG | 102 (12.4) | 11 (17.7) | 91 (12.0) | 10.78% |  |
| **rs11983224** |  |  |  |  |  |
| TT | 341 (41.4) | 23 (37.1) | 318 (41.8) | 6.74% | 0.266 |
| CT | 366 (44.5) | 26 (41.9) | 340 (44.7) | 7.10% |  |
| CC | 116 (14.1) | 13 (21.0) | 103 (13.5) | 11.21% |  |
| **rs7792678** |  |  |  |  | 0.463 |
| AA | 318 (38.6) | 24 (38.7) | 294 (38.6) | 7.55% |  |
| AC | 375 (45.6) | 25 (40.3) | 350 (46.0) | 6.67% |  |
| CC | 130 (15.8) | 13 (21.0) | 117 (15.4) | 10.00% |  |
| **rs7811849** |  |  |  |  | 0.463 |
| TT | 323 (39.2) | 24 (38.7) | 299 (39.3) | 7.43% |  |
| TA | 371 (45.1) | 25 (40.3) | 346 (45.5) | 6.74% |  |
| AA | 129 (15.7) | 13 (21.0) | 116 (15.2) | 10.08% |  |
| **rs1149515** |  |  |  |  |  |
| TT | 18 ( 2.2) | 2 ( 3.2) | 16 ( 2.1) | 11.11% | **0.019** |
| CT | 236 (28.7) | 27 (43.5) | 209 (27.5) | 11.44% |  |
| CC | 569 (69.1) | 33 (53.2) | 536 (70.4) | 5.80% |  |
| **rs191786** |  |  |  |  |  |
| TT | 40 ( 4.9) | 2 ( 3.2) | 38 ( 5.0) | 5.00% | **0.066** |
| CT | 287 (34.9) | 14 (22.6) | 273 (35.9) | 4.88% |  |
| CC | 496 (60.3) | 46 (74.2) | 450 (59.1) | 9.27% |  |

Cases, patients experiencing a MACE within one year after the PCI; controls, patients not experiencing MACE during the year following the PCI; N (%); P value from a chi-square test examining differences in MACE rate by genotypes.

